# Global genetic diversity patterns and transmissions of SARS-CoV-2

**DOI:** 10.1101/2020.05.05.20091413

**Authors:** Zhi-wei Chen, Zhao Li, Hu Li, Hong Ren, Peng Hu

## Abstract

**Background:** Since it was firstly discovered in China, the SARS-CoV-2 epidemic has caused a substantial health emergency and economic stress in the world. However, the global genetic diversity and transmissions are still unclear.

**Methods:** 3050 SARS-CoV-2 genome sequences were retrieved from GIASID database. After aligned by MAFFT, the mutation patterns were identified by phylogenetic tree analysis.

**Results:** We detected 17 high frequency (>6%) mutations in the 3050 sequences. Based on these mutations, we classed the SARS-CoV-2 into four main groups and 10 subgroups. We found that group A was mainly presented in Asia, group B was primarily detected in North America, group C was prevailingly appeared in Asia and Oceania and group D was principally emerged in Europe and Africa. Additionally, the distribution of these groups was different in age, but was similar in gender. Group A, group B1 and group C2 were declined over time, inversely, group B2, group C3 and group D were rising. At last, we found two apparent expansion stages (late Jan-2020 and late Feb-2020 to early Mar-2020, respectively). Notably, most of groups are quickly expanding, especially group D.

**Conclusions:** We classed the SARS-CoV-2 into four main groups and 10 subgroups based on different mutation patterns at first time. The distribution of the 10 subgroups was different in geography, time and age, but not in gender. Most of groups are rapidly expanding, especially group D. Therefore, we should attach importance to these genetic diversity patterns of SARS-CoV-2 and take more targeted measures to constrain its spread.

## Introduction

In December 2019, a novel coronavirus, severe acute respiratory syndrome coronavirus 2 (SARS-CoV-2), was first discovered in Wuhan, China^1^. The increasing epidemiological and clinical evidence implicates that the SARS-CoV-2 has lower pathogenicity than SARS-CoV but stronger transmission power^2^. To date, more than 1, 439, 516 people from 212 countries of six continents have been confirmed cases and more than 85, 711 have died from the rapidly-spreading SARS-CoV-2 (WHO, 2020)^3^. In addition, tens of thousands of new confirmed cases have been reported daily, which is now straining or overwhelming health care systems around the world^3^. SARS-CoV-2 is a betacoronavirus, which harbors a linear single-stranded positive RNA genome. The SARS-CoV-2 genome is about 30, 000 bp length, and contains 5’ untranslated regions (UTR), open reading frame (ORF)1ab gene encoding proteins for RNA replication, and other genes for non-structural proteins (nsp) and structural proteins. Like other RNA viruses, SARS-CoV-2 is prone to mutate due to the lack of proofreading function of RNA-dependent RNA polymerase (RdRp) in genome replication. These mutations may affect the virus’ gene function, virulence, transmission, detection and vaccine developing.

Hence, to know the genetic diversity of SARS-CoV-2 and how they spread in the world is important to control its epidemic. Recently, multiple studies have identified some high frequency mutations using SARS-CoV-2 sequences from GISAID database, such as 8782C>T, 23403A>G, 26144G>T and 28144T>C^4-11^. However, most of them were small sample size (less than 500 sequences) or only focus on part of genomes. The global genetic diversity of SARS-CoV-2, mutation patterns and transmissions are still unclear so far.

To answer these questions above, we analyzed 3050 SARS-CoV-2 genome sequences from GISAID database and found that based on different mutation patterns, the SARS-CoV-2 was classified into four main groups and 10 subgroups. The distribution of different groups of SARS-CoV-2 was varied in geography, time and age, but was similar in gender. Two apparent expansion stages were identified, notably, the second stage is still rapidly expanding in most of groups.

## Methods

### Genome sequences retrieval and alignment

Total 5848 sequences of the SARS-CoV-2 from the infected individuals were retrieved from the GISAID database^12^ (https://www.gisaid.org/) as of April 10, 2020. After screen, only the complete genomes of high-coverage were included in the dataset. The sample information including location countries, collection date, gender and age were retrieved from each sequence.

The complete genome sequences were aligned with the reference genome of SARS-CoV-2 (NC_045512) by MAFFT (version 7). The aligned genomes were then edited manually according to the reference sequence by BioEdit software. In the alignment, we found that the 5’UTR and 3’UTR contain missing and ambiguous sites in most of sequences, whereas, to largely preserve more sequence’s sites information, we just analyzed position 201-29674 (removed 3’UTR and part of 5’UTR regions) in the following analyses.

### Mutation analysis

Mutation was described as the replacement of the nucleotide in the reference sequence with one of A, T, G, C and gap. Degenerate bases were not considered as a mutation in this study. For ease to visualize the mutations, heatmap was drew by pheatmap package in R language. To investigate the relation between these mutations, DnaSP^13^ was used for linkage disequilibrium (LD) analysis. To better analysis mutation pattern of SARS-CoV-2, we performed a phylogenetic analysis. Average linkage (UPGMA) method with bootstrap value of 100 replicates was used for the construction of phylogenetic tree using MEGA X software. And then, tree was painted by iTOL^14^ (version 5).

#### Statistical analysis

All data were presented as rates (%) and analyzed statistically using the chi-squared test with SPSS 20 software (SPSS Inc., Chicago, IL, USA). p value ≤ 0.05 was considered statistically significant.

## Results

### Characteristics of SARS-CoV-2 sequences

Total 5848 sequences of the SARS-CoV-2 were retrieved from GISAID database at April 10, 2020. After screen, 3050 high-coverage SARS-CoV-2 strains were included in this study and listed in Table S1. The sample collection dates were from Dec 24, 2019 to April 6, 2020. The sample location covered all the six continents, involved 56 countries. Among them, 2195 (71.97%) sequences contained gender (1203 males and 992 females) and 2191 (71.84%) sequences contained age (range 0 to 102 year).

### Genomic diversity of SARS-CoV-2

In total, 1994 positions were detected mutations from all the 3050 sequences (Fig. S1). The results showed a large mutation diversity in these virus isolates. Most of them were found with low incidence (<1%). Only 46 positions occurred mutations with a >1% frequency, of them, 17 high frequency (>6%) mutation were identified. To ease know the global diversity of SARS-CoV-2, we used heatmap to visualize the 46 positions mutation, with time and geographic order (Fig. 1). In general, the mutation patterns were complicated and continuously evolving in the world. As shown in Fig. 1A, in Dec-2019, none of the 46 mutations were discovered. But with time goes on, more and more mutations were detected. In addition, we also found the mutation patterns were different between six continents. It seemly that mutations were relatively low in Asia, whereas more complicated in Europe. (Fig. 1B)

**Fig. 1.**
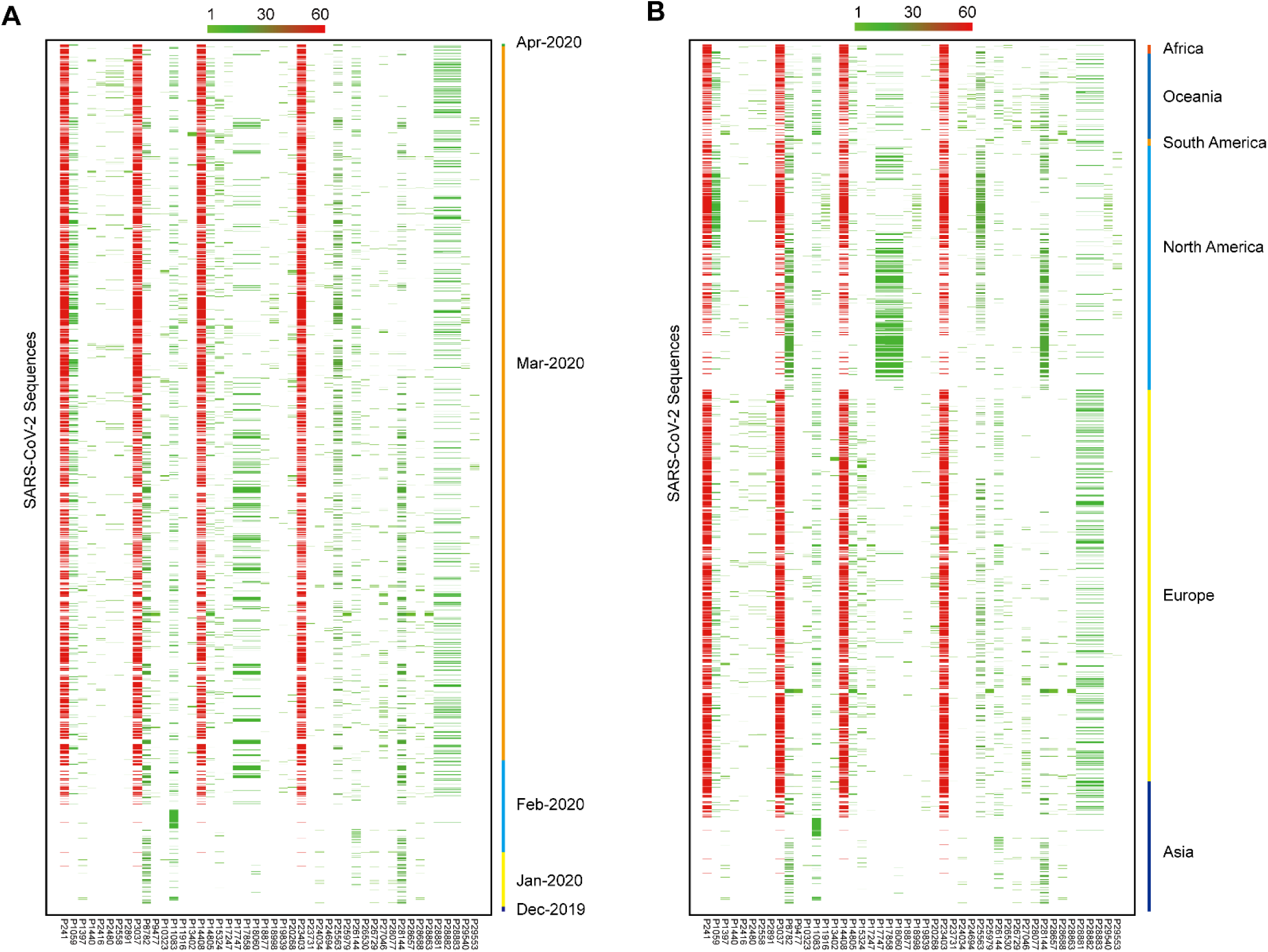
The distribution of mutations (>1%) of SARS-CoV-2. (A) The distribution of mutations (>1%) of the 3050 SARS-CoV-2 sequences over time. (B) The distribution of mutations (>1%) of the 3050 SARS-CoV-2 sequences in geography order. Green to Red means the frequency of mutations from 1% to 60%.

Further, we analyzed the 17 high frequency mutations in detail. We found that the mutations are not equally distributed. More than half of these mutations (nine mutations) presented in ORF1ab, followed by ORF9 (three mutations), ORF3a (two mutations), and at last only one mutation was detected in 5’UTR, ORF2 and ORF8, respectively. Of the 16 mutations in coding sequences, 11 mutations were non-synonymous mutations and five mutations were synonymous mutations. The most common mutation was 241C>T in 5’UTR (56.82%) and the most uncommon mutation was 14805C>T in ORF5 (8.59%). (Table 1)

**Table 1:** High-frequency (>6%) mutations in SARS-CoV-2 genome.

| nt mutation | Gene | Protein | AA mutation | Number | Frequency (%) |
| --- | --- | --- | --- | --- | --- |
| 241C>T | 5'UTR |  |  | 1733 | 56.82 |
| 1059C>T | ORF1ab | nsp2 | T85I | 514 | 16.85 |
| 3037C>T | ORF1ab | nsp3 | F105F | 1729 | 56.69 |
| 8782C>T | ORF1ab | nsp4 | S75S | 611 | 20.03 |
| 11083G>T | ORF1ab | nsp6 | L37F | 419 | 13.74 |
| 14408C>T | ORF1ab | nsp12 (RdRp) | P323L | 1709 | 56.03 |
| 14805C>T | ORF1ab | nsp12 (RdRp) | Y455Y | 262 | 8.59 |
| 17747C>T | ORF1ab | nsp13 (helicase) | P504L | 385 | 12.62 |
| 17858A>G | ORF1ab | nsp13 (helicase) | Y541C | 395 | 12.95 |
| 18060C>T | ORF1ab | nsp14 (3'-to-5'exonuclease) | L6L | 401 | 13.15 |
| 23403A>G | ORF2 (S) | spike glycoprotein | D614G | 1729 | 56.69 |
| 25563G>T | ORF3a | ORF3a protein | Q57H | 626 | 20.52 |
| 26144G>T | ORF3a | ORF3a protein | G251V | 274 | 8.98 |
| 28144T>C | ORF8 | ORF8 protein | L84S | 608 | 19.93 |
| 28881G>A | ORF9 (N) | nucleocapsid phosphoprotein | R203K | 491 | 16.10 |
| 28882G>A | ORF9 (N) | nucleocapsid phosphoprotein | R203R | 490 | 16.07 |
| 28883G>C | ORF9 (N) | nucleocapsid phosphoprotein | G204R | 490 | 16.07 |

Note: The mutation positions are on the reference genome. Nucleotide T represents nucleotide U in SARS-CoV-2 RNA virus genome. Red, non-synonymous mutation. RdRp, RNA-dependent RNA polymerase; nsp, non-structural proteins.

### Mutation patterns of SARS-CoV-2

Even though the mutations were complicated and evolving in the SARS-CoV-2 genome, we still tried to find some regular or patterns of these mutations. After linkage disequilibrium (LD) analysis, we did find some linkage mutations. The most common co-mutations were 241C>T, 3037C>T, 14408C>T and 23403A>G; followed by 8782C>T and 28144T>C; 17747C>T, 17858A>G and 18060C>T; 28881G>A, 28882G>A and 28883G>C (Table S2).

To better summarized these mutation patterns, we conducted a phylogenetic tree of all the 3050 SARS-CoV-2 genome sequences. As shown in Fig. 2A, based on the 17 high frequency mutations, these sequences were classed into four main groups (group A, B, C and D) and 10 subgroups (group A, group B1-B3, group C1-3 and group D1-3). And distinguished mutation patterns of each group were listed in Fig. 2B. Group A occurred none mutation in any of the 17 positions. Group B exhibited two primary mutations 8782C>T and 28144T>C. Group C contained mutations 11083G>T or/and 26144G>T. Group D included four main mutations 241C>T, 3037C>T, 14408C>T and 23403A>G.

**Fig. 2.**
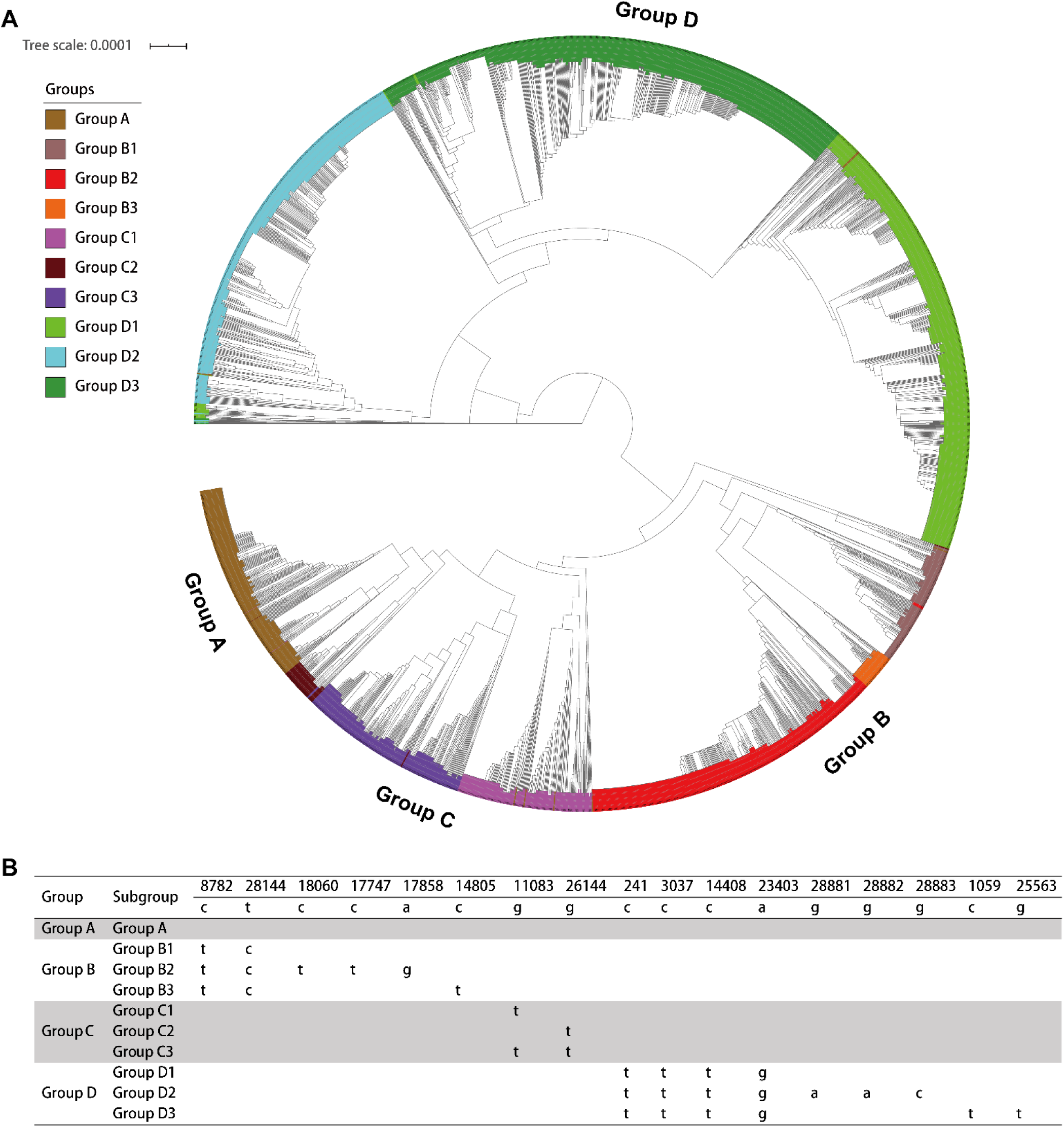
The mutation patterns of SARS-CoV-2. (A) 10 subgroups were classed by phylogenetic tree analysis of the 3050 SARS-CoV-2 genome sequences. (B) The detailed mutation patterns in different groups.

### The distribution of mutation patterns of SARS-CoV-2 in geography

Since we classified 3050 SARS-CoV-2 genome sequences into 10 subgroups mutation patterns, we want to know how these mutation patterns distribute in the world next.

In geography, there was significantly different of the 10 subgroups distributions between six continents (Fig. 3A). In detail, group A mainly presented in Asia and group D (including group D1-3) was mostly found in Europe and Oceania. Group B2 was primarily detected in North America, however, as an adjacent continent, South America was largely composed by group B3 and group D (including D1 and D2). At last, the main mutation pattern was group D (containing D1 and D3) in Africa.

**Fig. 3.**
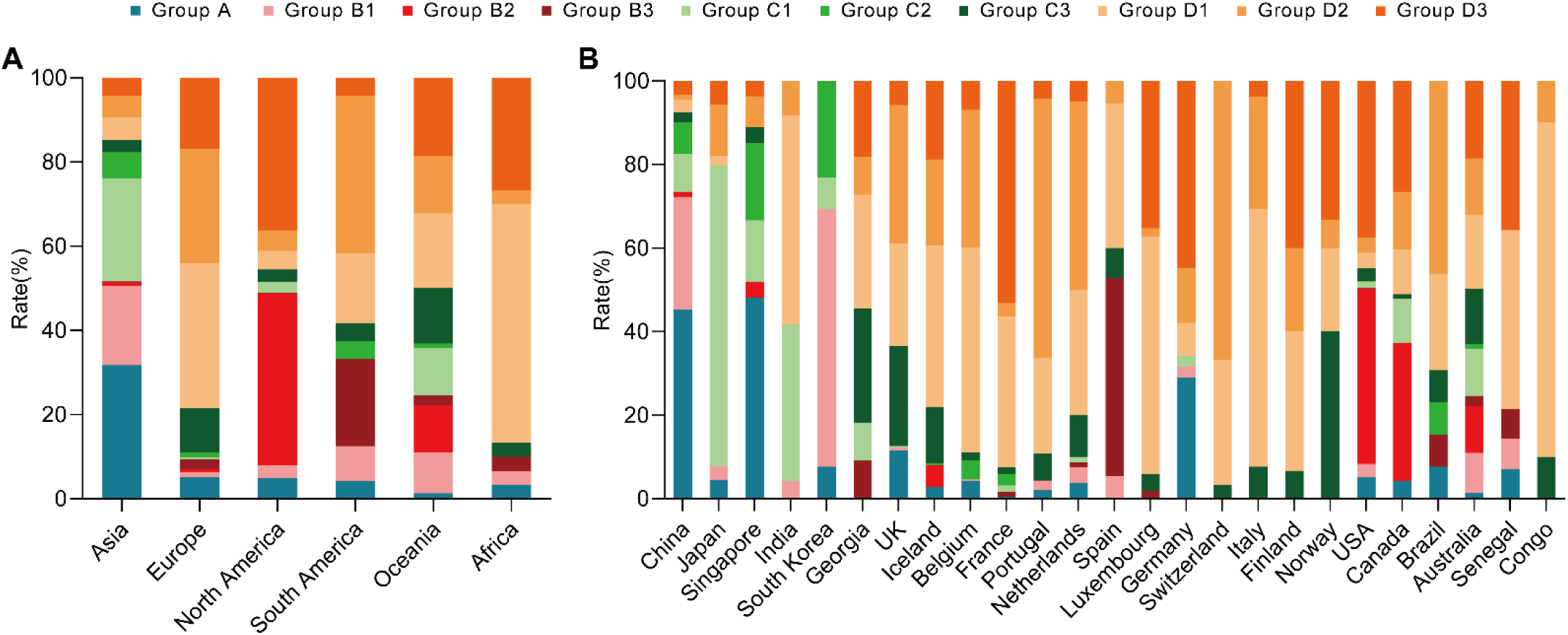
The distribution of the 10 subgroups mutation patterns in the world. (A) The distribution of the 10 subgroups mutation patterns in geography. (B) The distribution of the 10 subgroups mutation patterns in countries. Only country uploaded sequences more than 10 was exhibited in (B).

Further analysis in countries, more complex distributions were observed. Even in the same continent, the compose of 10 subgroups were different, such as, China, Japan, Singapore, India and South Korea in Asia. But group D, as a primary mutation pattern, was similarly discovered in all Europe countries (Iceland, France, United Kingdom, Netherlands, Belgium, Portugal, Switzerland, Spain, Germany, Italy, Finland and Luxembourg). (Fig. 3B)

### The distribution of mutation patterns of SARS-CoV-2 in gender and age

Even though not all sequences had genders and ages, there were still more than 2000 sequences for us to analyze whether gender and age have effect on the distribution of mutation patterns.

In gender, we found that most of groups were similar between males and females, except for group A was relatively low observed in females (Fig. S2A). Further analysis, we found this discrepancy was only presented in Europe (Fig. S2B).

In age, we found most of groups were presented a fluctuant trend with age. Interestingly, we found that group D2 was decreased, inversely, group D3 was increased with age. (Fig. S3A) This phenomenon was mainly observed in Europe and North America (Fig. S3B).

### The distribution of mutation patterns of SARS-CoV-2 over time

In total, the SARS-CoV-2 is continually evolving and the distribution of 10 subgroups mutation patterns are also dynamic with the time goes on. In Dec-2019, only group A was existed (100%), and then it declined in Jan-2020 (47.64%), in Feb-2020 (21.67%) and almost disappeared in Mar-2020 (3.52%). Similar with group A, group B1 and group C2 also shown decreased trend with time. On the contrary, group B2 firstly occurred in Jan-2020 (3.66%), increased step by step, and up to 14.22% in Mar-2020.

Analogical with group B2, we found that group D and group C3 (especially group D) also shown a fast expansion trend since their firstly emerged. The group C1 was different with other groups, which presented an increased trend in Feb-2020, whereas, sharply decline in Mar-2020. (Fig. 4A) Combined with the geographic data, more findings were observed. Firstly, the SARS-CoV-2 was first reported in Asia at Dec-2019, and transmitted to Europe, North America and Oceania at Jan-2020, then spread to South America and Africa at Mar-2020. (Fig. 4B). Secondly, in the early stage (Jan-2020), the distribution of 10 subgroups were similar between Asia and North America. And then, group A and group B1 decreased with time in both continents, group B2 increased in North America gradually, instead of, group C1 was scaling-up in Asia with time. (Fig. 4B)

**Fig. 4.**
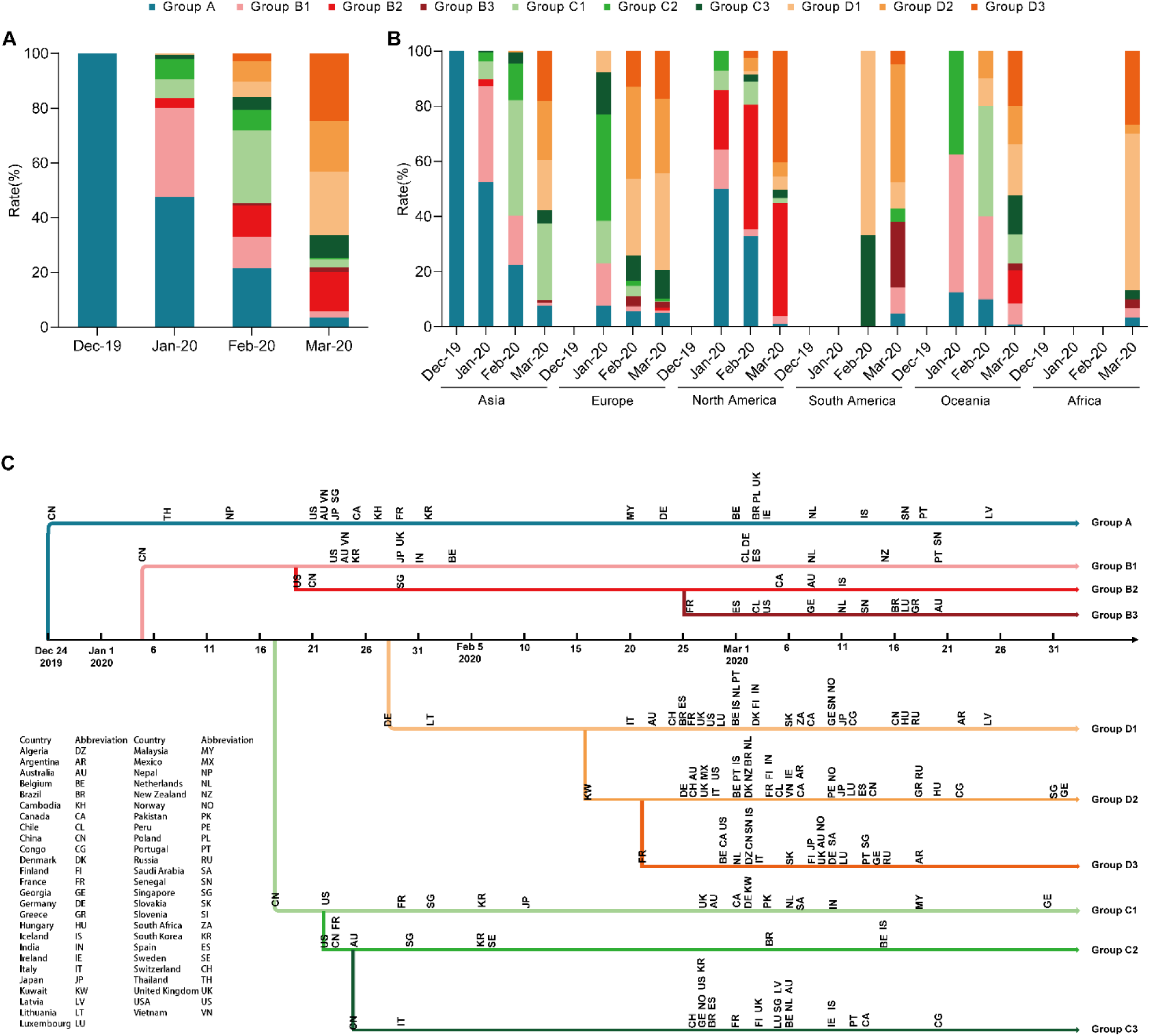
The distribution and transmissions of the 10 subgroups mutation patterns over time. (A) The distribution of the 10 subgroups mutation patterns over time in total. (B) The distribution of the 10 subgroups mutation patterns over time in geography. (C) The transmissions of the 10 subgroups mutation patterns between countries over time. Eight sequences in Apr-2020 and other 17 sequences containing only year, no specific time, were excluded for the analysis in (A) and (B). The country was only shown at the place where it was firstly appeared in each group in (C).

### The transmissions of mutation patterns of SARS-CoV-2 in the world

To comprehensively and systematically understand the different mutation patterns’ transmissions, we drew a flowchart using all available countries’ data in this study (Fig. 4C). In general, there were two fast global expansion stages in the 10 subgroups. First expansion stage was occurred at late Jan-2020, which contained group A, group B1, group C1 and group C2, which were firstly observed in China and USA. After a relatively stable stage in Feb-2020, another bigger expansion stage was emerged around the late Feb-2020 to early Mar-2020, which included all other 9 groups but group C2. This expansion stage was mainly started from Europe countries, especially in France and is still expanding with time. (Fig. 4C)

In detail, group A was the first observed in China at 2019/12/24, and started its first expansion to North America (USA, 2020/1/21) and Oceania (Australia, 2020/1/22), and second expansion to Europe countries and South America (Brazil, 2020/3/3). (Fig. 4C)

For group B, group B1 was also first observed in China at 2020/1/5, and the expansion pattern was similar with group A, just at a bit late. Before its first expansion, group B2 emerged in USA at 2020/1/19 for the first time, followed by China (2020/1/21), Singapore (2020/1/29), Canada (2020/3/5), Australia (2020/3/8) and Iceland (2020/3/11). At 2020/2/25, group B3 was firstly detected in France, and then quickly appeared in Europe, South America, North America, Africa and Australia. (Fig. 4C)

For group C, group C1 was firstly discovered in China at 2020/1/18, and then started the rapid expansion stage since 2020/2/22. Group C2 firstly emerged in USA at 2020/1/22, and started the first expansion immediately to the world. Group C3 was firstly appeared in China at 2020/1/25, after detected in Italy at 2020/1/29, it was stable during the Feb-2020 until 2020/2/26 started a quickly expansion. (Fig. 4C)

For group D, group D1 was firstly appeared in Germany at 2020/1/28, and then expanded quickly to other Europe countries since 2020/2/20. Group D2 was firstly observed in Kuwait at 2020/2/16 and group D3 was firstly detected in France at 2020/2/21. Both the groups expanded rapidly to Europe, Oceania and North America since their appeared. (Fig. 4C)

## Discussion

The SARS-CoV-2 epidemic has caused a substantial health emergency and economic stress in the world. Hence, understanding the global diversity and transmission of SARS-CoV-2 are critical in disease control. In this study, we used GIASID database to retrieve all available SARS-CoV-2 genome sequences, hoping to answer this question. First of all, we classed the global SARS-CoV-2 sequences into four main groups and 10 subgroups based on their different mutation patterns. Next, we found that these groups were markedly different in geography and time. This difference was also observed in age, but not in gender. At last, we systematically characterized the transmission of SARS-CoV-2 in the world and found two apparently expansions stages of the 10 subgroups with time. Notably, the second stage is still expanding in most groups (especially group D).

SARS-CoV-2, as an RNA virus, was prone to mutant. Since it was firstly discovered in Wuhan, China, more and more mutations were reported. In this study, after aligned the 3050 SARS-CoV-2 genome sequences, we identified 1994 positions occurred mutations, including 17 high frequency (>6%) mutations. In the 17 mutations, 14 mutations (241C>T, 3037C>T, 8782C>T, 11083G>T, 14408C>T, 17747C>T, 17858A>G, 18060C>T, 23403A>G, 26144G>T, 28144T>C, 28881G>A, 28882G>A and 28883G>C) had been reported by serval studies previously^4-11^. Of note, three novel mutations, 1059C>T (T85I in nsp2), 14805C>T (Y455Y in RdRp) and 25563G>T (Q57H in ORF3a) were firstly discovered in this study. However, the effects of the two new non-synonymous mutations on SARS-CoV-2 were still unclear.

Notably, the SARS-CoV-2 is evolving, since its firstly reported in the world^4,5^. To date, four main groups of mutation patterns of SARS-CoV-2 were identified in this study, based on the 17 high frequency mutations. Group A (included none of the 17 mutations) was the firstly discovered SARS-CoV-2 strain in the world which was emerged in Wuhan, China for the first time since Dec-2019^15^. Group B (contained mutations 8782C>T and 28144T>C) was in accord with Tang at al. discovered S type SARS-CoV-2 strain^4^ and GISAID database reported S clade of SARS-CoV-2 strain^16^, respectively. Group C (included mutations 11803G>T or/and 26144G>T) was similar with GISAID database reported V clade of SARS-CoV-2 strain^16^. However, the V clade in GISAID database was only distinguished by mutation 26144G>T. In this study, we found that joined 11803G>T was more proper to classify the SARS-CoV-2 strains. Group D (contained mutations 241C>T, 3037C>T, 14408C>T and 23403A>G) was in line with GISAID database reported G clade of SARS-CoV-2 strain^16^. Even though only one mutation 23403A>G was used to differentiate the G clade in GISAID database, the mutations 241C>T, 3037C>T, 14408C>T and 23403A>G were linkage in this study. So, we considered that they were identical.

Notably, to better understand the global diversity and transmissions of SARS-CoV-2, we further classed the SARS-CoV-2 strains into 10 subgroups by more detailed mutation patterns in this study for the first time. When add in geography, gender, age and time data of SARS-CoV-2 sequence, the answer was clearer and clearer.

Group A was the predominate strain at the beginning of globally spreading^17,18^, however, it was waned over time. Even though it transmitted to Europe, North America, South America and Oceania, we found that Group A was still mainly observed in Asia. When the group A declined, the group B1 emerged at 2020/1/5 in China. After short-time local expansion, it spread to USA, Australia and other countries.

However, there was a paradoxical phenomenon, that is, group B2 firstly occurred in USA at 2020/1/19, which was sooner than group A and group B transmitted into USA (2020/1/21 and 2020/1/23, respectively). The reason maybe that the sequences of group A and group B1 (emerged before group B2) in USA was not acquired or uploaded. Since group B2 was observed, it quickly became the primary strains in USA and then spread to China, Canada and Australia. Interestingly, when the group B2 emerged in China and Australia, it was almost not expanded. Whereas, in Canada, an adjacent country of USA, it was expanded at once. This hint us the different groups of SARS-CoV-2 may be limited in geography or race.

Group C was detected relatively low in the world, which was mainly presented in Asia and Oceania. Similar with group A and group B1, group C2 was decreased over time, inversely, group C3 was increased step by step. But the dynamic of group C1 was different with the two others, which had a big expansion at Feb-2020. This was because that many SARS-CoV-2 sequences from Diamond Princess cruise ship in Japan was uploaded at Feb-2020. This also was the reason why large compose of group C1 presented in Japan.

Group D was the latest occurred over time, however, once appeared, it apace expanded and had been the major strains now in the world, especially in Europe and Africa. Interestingly, we found that the group D2 were tended to decreased with age, however, the trend of group D3 was in the opposite direction. This indicated that the group D2 and group D3 were tightly with age.

In addition, we found two apparent expansion stages in the world of the SARS-CoV-2 spreading. One was from China and USA to the other countries at late Jan-2020, another was from Europe to the other countries at the late Feb-2020 to early Mar-2020. Notably, the second stage is expanding now. Hence, we still need to be on the alert and take active measures to contain the SARS-CoV-2 spreading.

Our study has some limitations due to the nature of the SARS-CoV-2 genome data. First, the sample collection dates may not reflect the actual infection date and not all infected countries uploaded the sequences timely to the GISAID database, so the mutation patterns transmission analysis is only approximate. Second, because some countries have not sequenced enough virus samples (such as Africa) or some countries uploaded samples from single-source (such as the Diamond Princess cruise ship in Japan), the mutation pattern may be biased in specific country or continent. Nevertheless, our study had been included the most complete available SARS-CoV-2 sequences to date. Whether the different mutation patterns of SARS-CoV-2 will result in biological and clinical differences remains to be determined.

In conclusion, we classed the SARS-CoV-2 into four main groups and 10 subgroups based on different mutation patterns for the first time. The distribution of the 10 subgroups was varied in geography, time and age, but not in gender. After two apparent expansion stages, most of groups are rapidly expanding, especially group D. Therefore, we should pay attention to these genetic diversity patterns of SARS-CoV-2 and take more targeted measures to control its spread.

## Data Availability

All data are available in the manuscript.

## Acknowledgements

We gratefully acknowledge the authors and originating and submitting laboratories of the sequences from GISAID’s EpiFlu (TM) Database on which this research is based. We are grateful to Trevor Bedford (GISAID) for providing instructions and advice on the database. A table of the contributors is available in Table S1.

## Author contributions

PH and HR conceived and designed the study. ZC and ZL retrieved sequences from GISAID database. ZC, ZL, HL and PH analyzed the data. ZC wrote the paper. PH and HR edited the paper. All authors agreed to be accountable for all aspects of the work and approved the final version of the manuscript.

## Financial Supports

This work was supported in part by the Kuanren clinical key disciplines construction program.

